# Availability and accessibility to latrine and handwashing facilities during working hours among “boda-boda” drivers in Mwanza city

**DOI:** 10.1101/2023.05.03.23289453

**Authors:** Namanya Basinda, Peter Chilipweli, Emmanuel B. Kisaka, Maxwell P. Shoo, Theckla Tupa, Marystella Z Vingson, Julieth Laizer, Eveline Konje, Anthony Kapesa, Dominica Morona

## Abstract

**Background:** Water access and sanitation are two of the most fundamental critical urban services upon which people in cities depend. Lack of these services may pose a major threat of disease especially in low- and middle-income countries. Mwanza City in Tanzania faces serious challenges regarding to sanitation services to the public and occupational on mobile bases such as Boda-boda. Boda-boda drivers are mobile and often come up into contact with people which might lead to the spread of diseases through the fecal-oral route. Thus, the aim of this study was to determine the level of accessibility and the barriers to sanitation and hygiene among the boda-boda drivers in Mwanza city.

**Methods:** This was a community based cross sectional study carried out in Nyamagana District that involved a randomly selected 196 participants. Data were collected using a pretested questionnaire. Data was be analyzed for frequency distribution, proportion and percentages for quantitative variables, mean ± SD.

**Results:** Almost a quarter (27.05%) had difficulties towards accessing latrines when they needed to use one. This was almost in parallel with 27.04% of the participants claiming latrines are not convenient in the community. Almost thirty percent (32.14%) were not satisfied with the availability of clean latrines, 67.35% claimed there are community laws or rules in place that make it more likely them to use a latrine every time they need to defecate or urinate. 84.69% showed great awareness towards realizing the importance of using latrines instead of the opposite.

**Conclusion:** Our results indicate that some of the Boda-boda drivers claims that latrines are not accessible and convenient but also habit has been a limiting factor which makes them to practice open defecation. Thus, there is inevitable need for the authority to build more clean latrines and run them as a business oriented low-cost facility i.e., pay per use. Together with making strict laws that enforces the use of latrine among the boda-boda drivers should be a priority.

## 1. Background

Water access and sanitation can be considered as two of the most fundamental critical urban services upon which the management of health and wellbeing of people in cities depend[2]. According to UNICEF and WHO, more than 2.4 billion people which is around 40% of the global population lack access to adequate sanitation and are forced to dispose of their excreta in unimproved and unsanitary conditions[3]. Further, 946 million people worldwide still practice open defecation[4]. All these pose a major threat of disease specifically communicable diseases such as diarrhea, malaria, typhoid and other waterborne diseases, as well as malnutrition. All of these have serious threat to the human beings and death as 7% of deaths in low- and middle-income countries are due to poor sanitation and hygiene[5]. Improving sanitation is known to reduce transmission of enteric pathogens and intestinal parasites, hence reducing morbidity and mortality, especially in children in both rural and urban areas of Tanzania. All these effects can hinder development in our country if taken for granted.

Open defecation (OD), which is human practice of defecating outside rather than into a toilet, increases human exposures to enteric pathogens[6]. People may choose fields, bushes, forests, ditches, streets, canals or other open space for defecation[7]. It is considered a major risk to children’s health and in all groups of ages and ultimately development. Around 25% of the Sub-Saharan Africa population lacks access to basic sanitation[8]. The practice of open defecation is much more common in rural areas of Sub-Saharan Africa with estimated 35% of rural households, in contrast to 8% of urban households[8]. Mwanza City faces serious challenges regarding to sanitation services to the public[1]. Several efforts have been made by the government and NGO’s, Example the TZS 245 billion invested by the government in 22nd February 2017 for the water and sanitation project which covered Mwanza city and three satellite towns of Magu, Lamadi and Misungwi [According to UNHABITAT.ORG], But despite these efforts, existing facilities have continued to deteriorate and have also failed to meet the demand of the equally increasing population in Mwanza. Although there is strong evidence about barriers to latrine and sanitation usage in Tanzania, mostly to the population accessing public latrines. Boda-boda drivers are marked to be in contact with many people per day which can easier transmit microbes is they are not hygienic and most of them use public latrines while at work, there is very little data that talk about the availability and access of latrines and sanitation to boda-boda drivers as for this study in Mwanza. They are mobile and often come up into contact with people which might lead to the spread of diseases. The primary objective of this study is to determine availability and access of good sanitation among the boda-boda drivers but the secondary objective is to determine factors which may hinder the effective accessibility and of the latrine during working hours among the Boda-boda drivers.

## 2. Materials and Methods

### 2.1. Study area

The study was conducted in Nyamagana District, in Mwanza. Mwanza is a port city on the shore of Lake Victoria, in northern Tanzania. Nyamagana District is one of the seven districts of the Mwanza Region of Tanzania. It is bordered to the north by Ilemela District, to the east by Magu District, to the south by Misungwi District and to the west by the Mwanza Bay of Lake Victoria. Part of the region’s capital, the town of Mwanza, is within Nyamagana District. According to Tanzania Census 2012 and National Bureau Statistics the population of the Nyamagana District was 363,452.

As of 2020 Mwanza city in general has a total number of approximately 18,500 boda-boda drivers. This is according to the U.W.P (Umoja wa Waendesha Pikipiki) Mwanza located near Mabatini.

### 2.2. Sample size, participants’ enrolment and data collection

#### 2.2.1. Sample size

The study involved “boda-boda” drivers at Nyamagana District, Mwanza City. A sample size of 196 Boda-boda drivers was estimated using fisher’s formula. The sample size was calculated from fisher’s formula [11].

#### 2.2.2. Inclusion criteria

The study involved any boda-boda driver, registered by the government for income purposes, above 18 years of age.

#### 2.2.3. Exclusion criteria

The study did not involve private boda-boda drivers who are not using their boda-boda for income purposes. The study also did not involve part time boda-boda drivers.

#### 2.2.4. Sampling procedure

The boda-boda drivers were recruited by a random sampling technique from the list obtained from their leaders, from Nyamagana district that fulfilled the inclusion and exclusion criteria, from 1st September to 30th September. The “vijiwe” (Vijiwe” – *is a Swahili word that means the place were the boda-boda drivers stay while waiting for passengers. It’s literally more like their stations or where you can find them*) were selected by random selection (*there are over 300 vijiwe in Nyamagana District alone according to U.W.P*.). Boda-boda drivers to participate in the research from the randomly selected vijiwe were also selected randomly from each Kijiwe (without forgetting the inclusion and exclusion criteria). With the help of the leaders in each “kijiwe” we explained to them about the research and its importance, and gave them questionnaires for filling.

### 2.3 Data collection

The data for the study was collected using structured questionnaires (which were in English and Swahili) in order to achieve the broad objective for the research and the specific objectives. The questionnaires had open -ended questions which were administered to the participants for self-filling by the data collectors who provided introduction and procedure on filling to the participants before giving them a tool. The tool was first tested to some various Vijiwe at Nata area where some Boda-boda are found.

### 2.3. Data analysis

Data collected was cleaned to exclude errors, re-organize, coded and processed for efficient analysis. Access to the data was limited to the researcher and the supervisor at the initial stage of the research untill completion. Data was also analyzed with Statistical package for Social Sciences (SPSS) version 22 and Microsoft Excel. Data was also analyzed for frequency distribution, proportion and percentages for quantitative variables, mean ± SD,

### 2.4. Ethics

The clearance to carry out this study was sought from the joint CUHAS and BMC Research, Ethic and Review Committee. Permission obtained from Regional and District Authorities. The benefits of the study were explained to participants, then voluntary informed consent signed by the participants. Confidentiality was ensured by keeping ensuring the participants no one will see their answers specifically by name. Example the questionnaires had no place where they write their names.

## 3. Results

### 3.1. Sociodemographic Characteristics of Respondents

A total of 196 male boda-boda drivers aged between 18-49 years were interviewed, with the median age of 27 years. Majority of respondents (83.67%) were either married or single. Majority of the respondents had an experience of less than 8 years.

4.08% did not attend school, 14.8% did not complete primary school, 28.06% completed primary school, 19.4% didn’t complete secondary school, 33.67% completed secondary school AND none attained higher level of education. Table 1 below which shows the sociodemographic information.

**Table 1.** Sociodemographic data.

| Age group | Frequency<br>(n=196) | Percentage (%) |
| --- | --- | --- |
| 11-20 | 19 | 9.69 |
| 21-30 | 122 | 62.24 |
| 31-40 | 43 | 21.94 |
| 41-50 | 12 | 6.12 |
| **Gender** |  |  |
| Male | 196 | 100 |
| Female | 0 | 0 |
| **Marital status** |  |  |
| Married | 97 | 49.49 |
| Co-habiting | 21 | 10.71 |
| Divorced | 11 | 5.62 |
| Widow | 0 | 0 |
| Single | 67 | 34.18 |
| **Level of education** |  |  |
| Not attended school | 8 | 4.08 |
| Not completed primary school | 29 | 14.8 |
| Completed primary school | 55 | 28.06 |
| Not completed secondary school | 38 | 19.4 |
| Completed secondary school | 66 | 33.67 |
| Higher education | 0 | 0 |
| **Did you use a latrine last day at work?** |  |  |
| Yes | 139 | 70.92 |
| No | 40 | 20.41 |
| Don't remember | 17 | 8.67 |
| **Latrine use habit** |  |  |
| Every time | 61 | 31.12 |
| Most of the time | 70 | 35.71 |
| Moderately | 42 | 21.43 |
| Not much | 20 | 10.2 |
| Not at all | 3 | 1.53 |

### 3.2. Availability and use of Latrine

Despite majority of the respondents (70.92%) admitting to using latrine the last time they went to urinate or defecate, only 31.12% admitted to having a habit of using the latrine every time they needed to. The results also showed that 35.71% admitted to using the latrine most of the time, 21.43% admitted using moderately, 10.2% admitted not much while 1.53% admitted not at all. Table 1 above which show demographic infromation.

### 3.3. Latrine use and cleanliness

On assessing the use latrine against cleanliness status of the latrine majority of the participants were convinced to use the latrine due to it cleanliness (48) but few use very clean latrine since it has some cost implication on the use (18) although most of the larine were normal in the sense that there were not that much clean and were free to use with no cost implications which made most participants to use. Thus, the use of latrine is highly promoted by its status of cleanliness and cost implication Figure 1 below.

**Figure 1.**
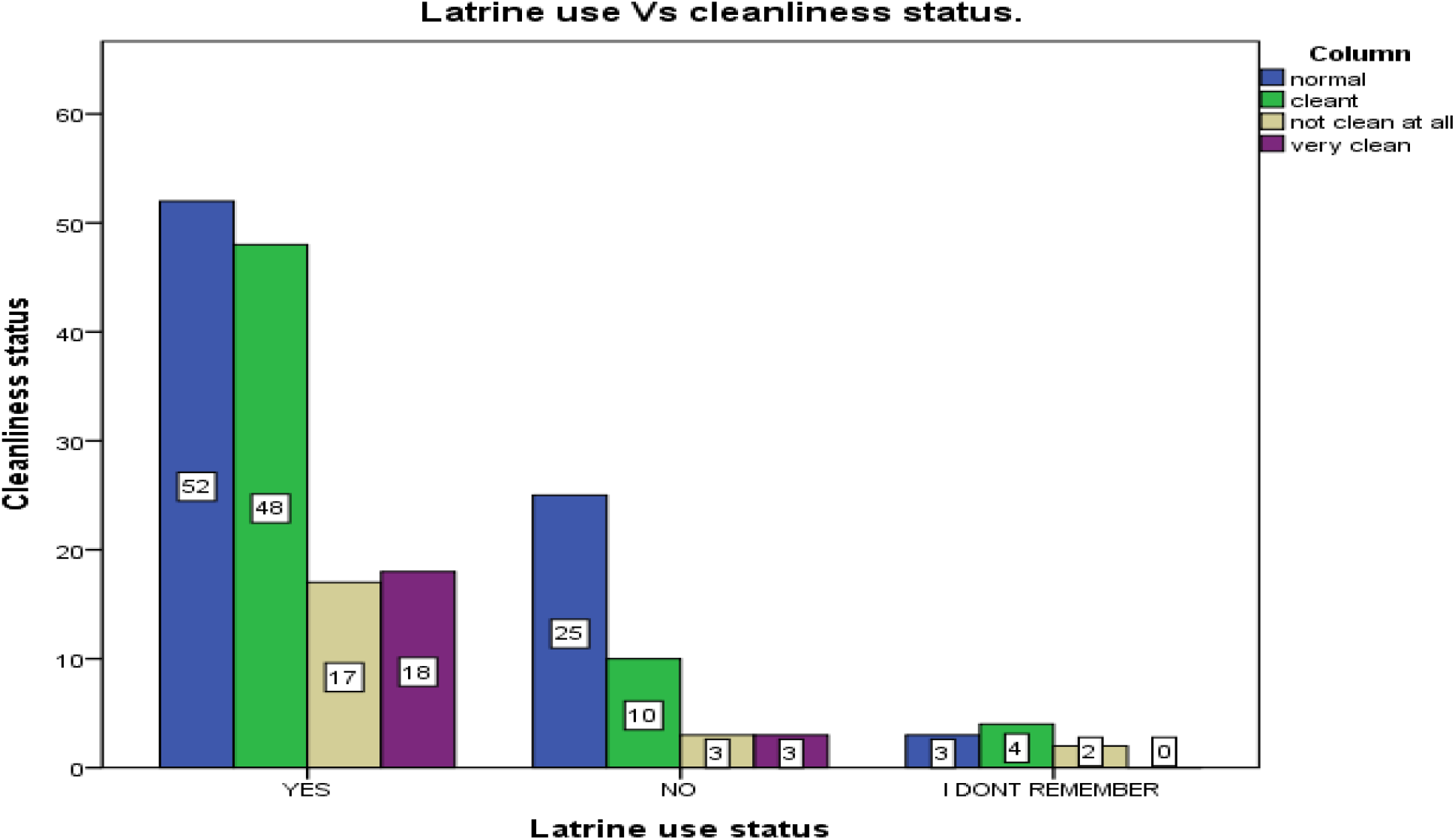
Latrine use and the Cleanliness. (*X- latrine use status, y-cleanliness frequency of the latrine)*

### 3.4. Latrine use and period of working

When latrine use and the period of working were compared it was seen that those working for two to one year had never and rarely use the latrine. This implies that Time is not a tangible aspect to justify that one will use the latrine Figure 2 below.

**Figure 2.**
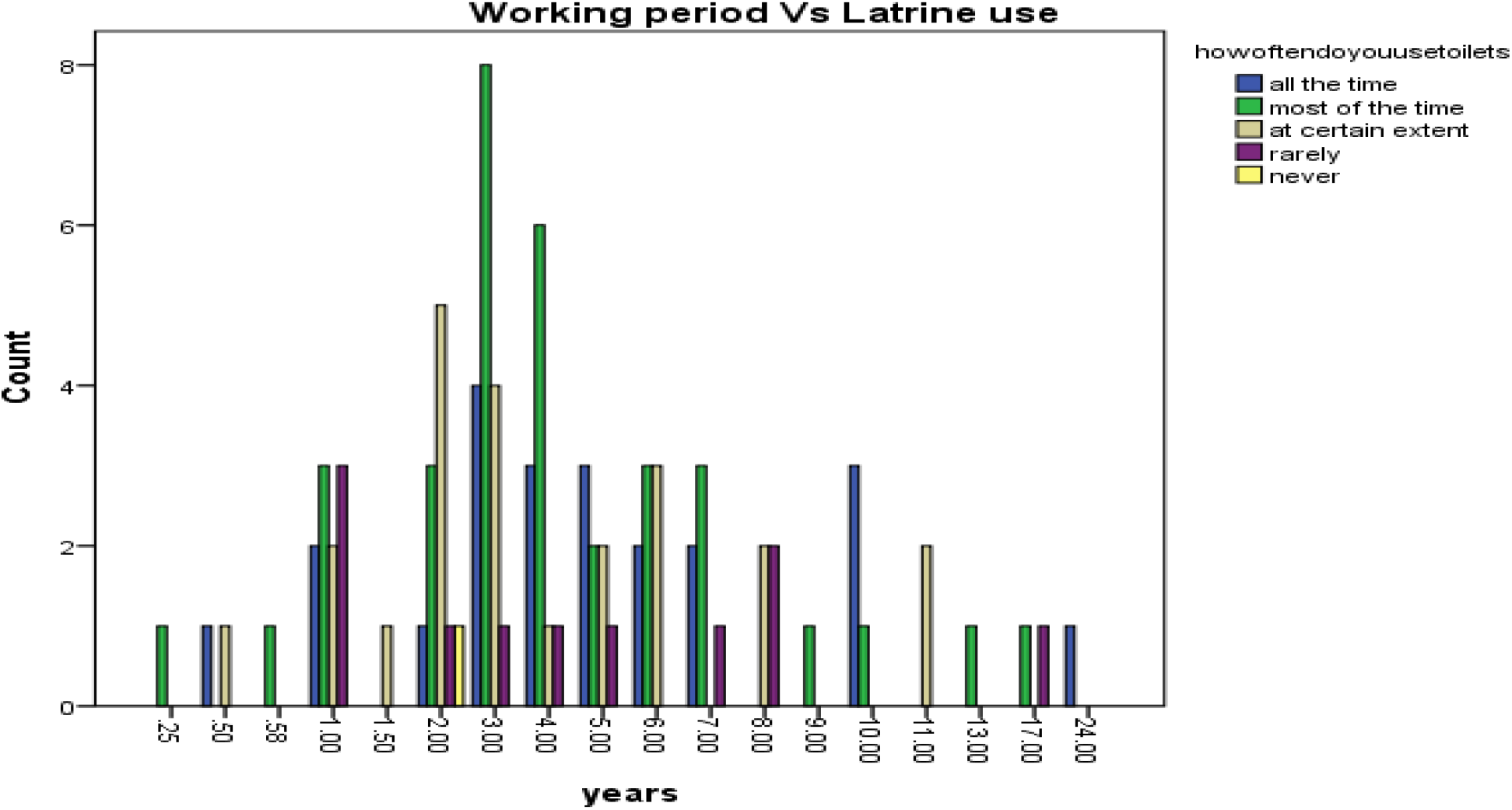
Latrine use and the working period of a “Boda-boda “ Worker. *(y- latrine use frequency, x- number of years working)*

### 3.5. Attributes for latrine use

#### 3.5.1. Accessibility and Convenience of Latrines

Nearly above a quarter of the participants (27.05%) had difficulties towards accessing latrines when they needed to use one. This was almost in parallel with 27.04% of the participants claiming latrines are not convenient in the community. Table 2 which show important attributes. Quarter of the population (25.51%) reported not getting any difficulties towards accessing latrines with 28.06% (that is almost one third of the population) claiming latrines are convenient.

**Table 2.** Accessibility and Convenience, Satisfaction with Availability of clean latrines and Affordability.

|  | Frequency<br>(n=196) | Percentage (%) |
| --- | --- | --- |
| **Difficulties accessing latrine** |  |  |
| Extremely difficult | 25 | 12.76 |
| Very difficult | 28 | 14.29 |
| Somewhat normal to find one | 93 | 47.45 |
| Not difficult | 28 | 14.29 |
| Not difficult at all | 22 | 11.22 |
| **Latrines convenience** |  |  |
| Extremely convenient | 20 | 10.2 |
| Very much | 35 | 17.86 |
| A moderate amount | 88 | 44.9 |
| Not much | 38 | 19.39 |
| Not convenient at all | 15 | 7.65 |
| **Satisfaction with latrines** |  |  |
| Very satisfied | 17 | 4.6 |
| Satisfied | 46 | 18.88 |
| Moderately | 87 | 44.39 |
| Dissatisfied | 37 | 23.47 |
| Very dissatisfied | 9 | 8.67 |
| **Any financial difficulties towards latrine access?** |  |  |
| Not at all |  |  |
| Not much | 9 | 4.6 |
| A moderate amount | 23 | 11.73 |
| Very much | 121 | 61.73 |
| An extreme amount | 29 | 14.8 |
|  | 14 | 7.14 |

#### 3.5.2. Satisfaction with Availability of clean latrines

About one third of the respondents (32.14%) were not satisfied with the availability of clean latrines while 23.48% of the respondents reported of being satisfied with their availability. But 44.39 were moderately satisfied with the availability of clean latrines. Table 2 which show Satisfaction with Availability of clean latrines.

### 3.6. Affordability

Only 21.94% of the respondents reported of having financial difficulties towards latrine access while only 16.33% reporting of not having any financial difficulties towards latrine access. Table 2. However, more than half of the respondents (61.73%) reported of having just a normal financial situation towards accessing latrines when they need to.

### 3.7. Product Attributes

Regarding product attributes, less than half of the participants (44.39%) reported that latrines are clean while only 10.2% claiming latrines are not clean at all.

Clearly a very large proportion of the respondents reaching 91.32 % acknowledged the hygienic nature of latrines, while the remaining percent (8.68%) lacking the knowledge that using latrines is always hygienic. Table 3 which show Product Attributes and cleanliness.

**Table 3.** Product Attributes and cleanliness.

|  | Frequency(n=196) | Percentage (%) |
| --- | --- | --- |
| **Cleanliness of latrines** |  |  |
| Not clean at all | 20 | 10.2 |
| Just normal (Moderate amount) | 89 | 45.4 |
| Very Clean | 64 | 32.65 |
| Extremely Clean | 23 | 11.74 |
| **Are latrines hygienic?** |  |  |
| I don't believe at all | 3 | 1.53 |
| Somewhat hygienic | 0 | 0 |
| Moderately | 14 | 7.14 |
| Mostly hygienic | 59 | 30.1 |
| Extremely hygienic | 120 | 61.22 |
| **Laws or rules that make it likely for a you to use a latrine** |  |  |
| Yes | 132 | 67.35 |
| No | 58 | 29.6 |
| Don't know | 6 | 3.06 |
| **Cultural rules or taboos that restrict one to use latrine** |  |  |
| Yes | 0 | 0 |
| No | 196 | 100 |
| Don't know | 0 | 0 |
| **How happy are you with access to sanitation information?** |  |  |
| Not happy at all | 30 | 15.3 |
| Somewhat unhappy | 55 | 28.06 |
| Very happy | 91 | 46.43 |
| Extremely happy | 20 | 10.2 |
| **Normality towards latrine use in the community** |  |  |
| Very normal | 17 | 8.67 |
| Somewhat Normal | 38 | 19.39 |
| Not normal | 98 | 50 |
| Not normal at all | 43 | 21.93 |

### 3.8. Social Support

Again, regarding social support from the government, more than half of the respondents (67.35%) claimed there are community laws or rules in place that make it more likely them to use a latrine every time they need to defecate or urinate WHILE 29.6% denying such a thing.NO ONE claimed that there are cultural rules or taboos that restrict them from using a latrine. More than half of the population (56.63%) were happy with access and availability to sanitation information, and the remaining percent (43.37%) were not happy. Table 3 which show Product Attributes and cleanliness.

### 3.9. Social Norms

Exactly 50% claimed it’s not a normal practice in the community to practice open defecation or urinating outside while 21.93% claimed it’s not a normal practice at all. Only a few proportions of the respondents (8.67%) claimed open defecation or urinating outside is a very normal practice in their community and the remaining percent (19.39%) claimed it’s somewhat normal. Table 3 which show Product Attributes and cleanliness.

### 3.10. Safety of and Health of Latrines

Majority of the respondents (84.69%) showed great awareness towards realizing the importance of using latrines instead of the opposite. Majority of the respondents (71.43%) claimed it would be very serious if him or his fellow boda-boda member gets a diarrhea disease, while 22.96% claimed it would be somewhat serious, 2.25% claimed it would not be serious at all. Table 4 which show distribution Table on Safety of and Health of Latrines and believes.

**Table 4.** Distribution Table on Safety of and Health of Latrines and believes.

|  | Frequency<br>(n=196) | Percentage |
| --- | --- | --- |
| **Believe latrines are safe to use?** |  |  |
| Never safe |  |  |
| Rarely safe | 0 | 0 |
| Some of the time safe | 3 | 1.53 |
| Most of the time | 27 | 13.78 |
| Always safe | 57 | 29.08 |
|  | 109 | 55.61 |
| **Possibility of getting diarrhea in the next 3months** |  |  |
| Very likely |  |  |
| Somewhat likely | 34 | 17.35 |
| Not likely at all | 64 | 32.65 |
| Don't know or won't say | 53 | 27.04 |
|  | 45 | 22.96 |
| **How serious would it be if you got a diarrhea disease?** |  |  |
| Very serious |  |  |
| Somewhat serious | 140 | 71.43 |
| Not serious at all | 45 | 22.96 |
| Don't know or won't say | 5 | 2.25 |
|  | 6 | 3.06 |
| **With your present knowledge, resources, and skills, could you use a latrine every time you needed to?** |  |  |
| Yes |  |  |
| Possibly | 115 | 58.67 |
| No | 62 | 31.63 |
| Don't know | 19 | 9.69 |
|  | 0 | 0 |
| **Difficulty towards remembering using a latrine every time you need to.** |  |  |
| Very difficult | 11 | 5.6 |
| Somewhat difficult | 45 | 22.96 |
| Not difficult at all. | 140 | 71.43 |
| Don't Know / Won't say | 0 | 0 |

### 3.11. Ability and Difficulty remembering to use a Latrine

More than a half of the respondents (58.67%) reported of having great ability towards using a latrine every time they need to, due to their knowledge, resources, and skills that they have. 31.63% said they could possibly do that while 9.69% said they could not. Table 4. Regarding difficulty remembering to use latrines, a large proportion of people (71.43%) reported it was not difficult at all to remember using a latrine. 22.96 reported it was somewhat difficult while only 5.6% had difficulties remembering. This showed that boda-bodas not remembering to use latrines wasn’t a big concern

## 4. Discussion

### 4.1. Latrine Use

Tanzania is estimated that 93 % of the population has access to a latrine. However, when assessing access to improved sanitation that figures drop to 24 %, depending on the definition of improved sanitation used [20].On our findings majority of the respondents admitting using latrine the last time they went to urinate or defecate, few admitted having a habit of using the latrine every time they needed to. This shows a large proportion of them of not having a habit of always using the latrines. A study by Tumwine [12] has shown that people living in the urban areas have a tendence of using latrine more as compared to rural since urban is endowed with more latrine which is the same from our findings.

But Beshonge [13] and Chagu[15] pointed on the challenges which makes most of the urban habitants not to use latrines such as lack of financial resources, rapid population increase, socioeconomic disparities among the urban inhabitants, topography, lack of skilled, and experienced personnel, inadequate policies and strategies, and people’s behaviors and attitudes. But also Chilipweli [23] showed behavior as the main challenge for latrine accessibility in the community which is almost the same from our findings which also pointed attitude in terms of habit as the constrains of using the latrine among Boda-boda drivers who are mostly predisposed in the urban settings.

Through the habit of not using latrines regularly, has negative impacts on the health and social wellbeing of communities, the environment and on the economy of the country at last. Lack of proper sanitation and hygiene predisposes the inhabitants to particularly poor health, high incidence of communicable diseases such as diarrhea, malaria, typhoid and other waterborne diseases, as well as malnutrition[9,18].As in Tanzania health burden due to poor sanitation and hygiene is significant, Diarrhea in the preceding two weeks is reported on average in 15 % of children under five years of age and results in 9 % of all mortality for this age group, Cholera and Typhoid is endemic in some areas of Tanzania and outbreaks are common [20].

A study in Ethiopia [19] showed presence of any latrine was related to greater awareness about importance to health to the community which is contrary with our findings which have shown that among those having the latrine few of them use them when they need to in the urban settings which indicate that the community is not aware on the health importance of using the latrine properly when needed.

Also our findings number of working years of the Boda-boda driver is not related with latrine use and awareness on its outcomes. This is contrary to Malik study [24] done in urban settings which showed knowledge on the latrine use of the respondent was much related with working status and number of years, level of education and family size, but through there was a clear gap between knowledge and actual practices. Thus practice is the main aspect in the use of latrine as compared to the knowledge participants have, so much effects have to be used on making the practice actual.

A combination of household-centered environmental sanitation (HCES) and community-led total sanitation (CLTS), two field-tested methodologies, has the potential to improve the sustainability of sanitation service interventions in the urban settings [14], thus for our settings it is important to make Boda-boda drivers as part of the implementation of the two proved effective methods for behavior change.

The government should build more clean latrines and run them as a business oriented low-cost facility i.e., pay per use. Together with making strict laws that enforces latrine use among the boda-boda drivers should also be a priority.

### 4.2. Accessibility and Convenience of Latrines

Access to improved sanitation is a key preventive measure against sanitary-related gastro-enteric diseases such as diarrhea. The sustainable Development Goal (SDG) 6, portrays on the aspect of providing safe water and sanitation for all, that the international community and governments must make WASH accessible and inclusive. But in low-income countries, only 1 in 3 people have access to basic hygiene services making them especially vulnerable [21]. On our findings almost one third of the population had difficulties towards accessing latrines when they needed to use them which was almost in parallel with participants claiming latrines are not convenient in the community. All this implies that, there is a lack of latrines which can be accessible in terms of Quality of the infrastructure, design, use, circulation inside while use, affordability and with safe water which could have big consequences to the health, social wellbeing and the economy of the country.

As we all know an unhealthy society is a loss of manpower in building the country, hence poor development. This raises the necessity of the government and the community at large in building more latrines so that people can access them more easily when they need to. Water provision should be insured to most of the public latrine and there should be a considerable bill for water in the public latrine as compared to other unit of use.

### 4.3. Satisfaction with Availability of clean latrines

On a study conducted in Kampala among slum dwellers by Tumwebazea (22) it shows that more than half of the respondents (51.7%) were not satisfied with their sanitation facilities whereby the determinants for satisfaction with the facilities used included the nature and type of toilet facilities used, their cleanliness, and the number of families sharing them. But on our findings more than one third of the respondents were not satisfied with the availability of clean latrines due to various factors such as its accessibility and other enabling factors.

This lack of satisfaction always discourage someone from using the toilet and thus leads to spread of diseases as mentioned earlier. This number is significant in raising awareness to the importance of not only having just latrines, but clean latrines so as to reduce greatly the incidence of communicable diseases that are its major consequence from affecting majority of the people in our society.

### 4.4. Affordability

The study pointed that the use of latrine is highly promoted by its status of cleanliness and cost implication, thus cost is one of the important factors which makes the latrine services either affordable or not. As it is having been observed that more than half of the respondents reported of having just a normal financial situation towards accessing latrines when they need to. However only 21.94% of the respondents claiming of having financial difficulties towards latrine access. This still shows there is a financial problem in the society. Boda-boda drivers (even most people in the society) who have a low income, spend it primarily on food and goods, with other items given low priority. It is difficult to convince these people to use their limited finances on sanitation when they have lived their entire life without it. Even when they are convinced that sanitation will be beneficially, the perceived high cost of installation keeps many people from adopting latrines[10]. There is inevitable need to make sure that latrine services are provided for free or with a cost which easy to be achieved by most of the community.

### 4.5. Product Attributes

Only few of the respondent’s claimed latrines are not clean at all. Clearly a very large proportion of the respondents reaching 91.32 % acknowledged that latrines should always be clean. Having unclean sanitation facilities leads to poor health, high incidence of communicable diseases such as diarrhea, malaria, typhoid and other waterborne diseases, malnutrition which impacts the economy of the country as explained before.

### 4.6. Social Support

Just over a quarter denied of presence of community laws or rules in place that make it more likely them to use a latrine every time they need to defecate or urinate. This data was in conjugation with 43.37% of the population who were not happy with access and availability to sanitation information. Presence of laws and legislations will help improve utilization of latrines because people won’t afford to lose time and money on something which they know they are not supposed to do according to the law as they could be arrested with it. This is more like saying the government needs to make laws stricter, that will increase chances of someone to use latrines.

### 4.7. Limitation

Disclosure of sensitive information from the participants to admit poor socially recognizable attitudes. Example using the bush to defecate for a grown man, may seem irrational and loss of awareness hence loss of respect to the candidate. To solve this, I tried to explain to the participant the importance of this study to the health of the community and development.

## 5. CONCLUSION

There is a general lack of persistent use of sanitation and hygiene facilities among boda-boda in Mwanza city. About a quarter (27%) have high difficulties in accessing sanitation and hygiene facilities. About 44.39% reported that latrines are clean while only 10.2% claiming latrines are not clean at all. Regarding social support from the government, almost one third (29.6%) denied the existence of community laws or rules that would make it more likely for them to use a latrine every time they need to defecate or urinate. This just shows how much more education is needed in the community and enforcement of the laws to encourage them to use latrines.

## Data Availability

data can be shared once there is necessity of doing so

## RECOMMENDATIONS

Most of the public areas had latrine but there were some attribute which makes most of the Boda-boda drivers not to use latrine provided such as convenience of the latrine, knowledge on the importance of using latrine, cleanliness, community rules and cost which they have to pay once used. Thus there is inevitable need to make sure that latrine services are provided for free or with a cost which easy to be achieved by most of the community especially occupations on mobile like Boda-boda and cleanliness on the public latrine should be improved.

With the leave no one behind goal our policy and strategy should also point Boda-boda as the important population to be addressed with WASH intervention as a separate group which also has a potential entity of causing infection and microbes spreading to the community.

